# SARS-CoV-2 antibody immunoassays in serial samples reveal earlier seroconversion in acutely ill COVID-19 patients developing ARDS

**DOI:** 10.1101/2021.02.15.21250916

**Authors:** Marie-Luise Buchholtz, Florian M. Arend, Peter Eichhorn, Michael Weigand, Alisa Kleinhempel, Kurt Häusler, Mathias Bruegel, Lesca M. Holdt, Daniel Teupser

## Abstract

**Objectives:** During the COVID-19 pandemic, SARS-CoV-2 antibody testing has been suggested for (1) screening populations for disease prevalence, (2) diagnostics, and (3) guiding therapeutic applications. Here, we conducted a detailed clinical evaluation of four Anti-SARS-CoV-2 immunoassays in samples from acutely ill COVID-19 patients and in two negative cohorts.

**Methods:** 443 serum specimens from serial sampling of 29 COVID-19 patients were used to determine clinical sensitivities. Patients were stratified for the presence of acute respiratory distress syndrome (ARDS). Individual serum specimens from a pre-COVID-19 cohort of 238 healthy subjects and from a PCR-negative clinical cohort of 257 patients were used to determine clinical specificities. All samples were measured side-by-side with the Anti-SARS-CoV-2-ELISA (IgG), Anti-SARS-CoV-2-ELISA (IgA) and Anti-SARS-CoV-2-NCP-ELISA (IgG) (Euroimmun AG, Lübeck, Germany) and the Elecsys Anti-SARS-CoV-2 ECLIA (Roche Diagnostics International, Rotkreuz, Switzerland).

**Results:** Median seroconversion occurred earlier in ARDS patients (8-9 days) than in non-ARDS patients (11-17 days), except for EUR N-IgG. Rates of positivity and mean signal ratios in the ARDS group were significantly higher than in the non-ARDS group. Sensitivities between the four tested immunoassays were equivalent. In the set of negative samples, the specificity of the Anti-SARS-CoV-2-ELISA (IgA) was lower (93.9 %) compared to all other assays (≥98.8 %) and the specificity of Anti-SARS-CoV-2-NCP-ELISA (IgG) was lower (98.8 %) than that of Elecsys Anti-SARS-CoV-2 (100 %).

**Conclusions:** Serial sampling in COVID-19 patients revealed earlier seroconversion and higher signal ratios of SARS-CoV-2 antibodies as a potential risk marker for the development of ARDS, suggesting a utility for antibody testing in acutely diseased patients.

## Introduction

Since the beginning of 2020, a large number of serological tests for antibodies against severe acute respiratory syndrome coronavirus 2 (SARS-CoV-2), the causative agent of coronavirus disease 2019 (COVID-19), has flooded the market to complement direct virus detection by PCR. As recommended by the Centers for Disease Control and Prevention, direct virus detection by PCR is essential and indispensable in acute diagnostics [1]. In contrast, the role of serological testing for antibodies against SARS-CoV-2 is less clear. It has been reported that median seroconversion occurs at 7-14 days [2-6], and later than PCR-positivity. In addition, it has been noted that individuals with mild or asymptomatic disease may only present delayed and transient serum titers of SARS-CoV-2 specific antibodies [7, 8]. This makes serological testing unsuitable for diagnostics in the early phase of disease. Nevertheless, SARS-CoV-2 serology may still play a role in diagnostics of patients suspected for a previous contact with SARS-CoV-2 and (false) negative PCR [6, 9, 10].

In contrast to diagnostics, it is without question that SARS-CoV-2 antibody testing has an important part in epidemiological studies. In these scenarios, the highest possible specificity of tests is of utmost importance, since the prevalence of SARS-CoV-2 is currently low in most populations, and therefore, only highly specific tests lead to acceptable false positive rates [11-14]. SARS-CoV-2 antibody testing may also be suitable to identify convalescent individuals for plasma donation and to identify potential vaccination responses, even though little is currently known about the protective effects of different types of SARS-CoV-2 antibodies [9, 15].

Main antigens to induce an immune response in the host with subsequent antibody production are the nucleocapsid (N) protein and the spike (S) protein with its receptor binding domain (RBD) [16]. Several Anti-SARS-CoV-2 immunoassays detect the N-protein, others the entire spike protein, its S1 subunit or the RBD, which is responsible for the entry of SARS-CoV-2 into the host cells via the ACE-receptor [17, 18]. Designing immunoassays with high specificities is challenging, given the homology of SARS-CoV-2 to other coronaviruses [2, 16, 18, 19]. Cross-reactivity may be observed with SARS-CoV and MERS-CoV, due to partial conservation of subunits of the S- and N-proteins [2, 19].

In the present study, we examined the performance of four CE-certified immunoassays detecting antibodies against the N- and the S1-proteins, two of which have received emergency use authorization by the U.S. Food and Drug Administration (FDA). These immunoassays can be automated and are suitable for rapid diagnostics in clinical routine. The two FDA approved tests were the Euroimmun Anti-SARS-CoV-2-ELISA (IgG) (EUR S-IgG) (catalog number: EI 2606-9601 G) and the Elecsys Anti-SARS-CoV-2 electrochemiluminescence immunoassay (Roche-Ab) by Roche (catalog number: REF 09203079190). These tests were complemented by the Euroimmun Anti-SARS-CoV-2-ELISA (IgA) (EUR S-IgA) (catalog number: EI 2606-9601 A) and the Euroimmun Anti-SARS-CoV-2-NCP-ELISA (IgG) (EUR N-IgG) (catalog number: EI 2606-9601-2 G) immunoassays. The EUR S-IgG and EUR S-IgA immunoassays detect IgG and IgA antibodies against the recombinant S1 domain of the SARS-CoV-2 spike protein, respectively. EUR N-IgG detects IgG-antibodies against a modified nucleocapsid protein and Roche-Ab detects antibodies (including IgG) against a renatured chaperone nucleocapsid fusion protein.

An important current clinical question in the COVID-19 pandemic is the early identification of patients with a high risk for severe clinical symptoms. Acute respiratory distress syndrome (ARDS) is a typical complication of COVID-19, frequently requiring therapy with ventilators [20]. Previous studies explored the question whether there is a correlation with the dynamics and level of SARS-CoV-2 antibody titers and the severity of COVID-19. Some studies reported an association of antibody titers with disease severity [3, 4, 21-23]. Previous studies were mainly based on cumulative samples of different individuals at different times over the course of disease. It has been proposed that sensitivity would be ideally determined at various days postsymptom onset [12]. In our study, we therefore used serial samples in hospitalized patients to assess immunoassay sensitivities after the onset of symptoms, where samples for a follow-up of at least 15 days were available.

Our study had two main objectives: (1) To evaluate antibody dynamics and sensitivities in serial samples from acutely ill COVID-19 patients with the stratification of the cohort in non-ARDS and ARDS patients, and (2) to assess the specificity of the four tests, side by side in a pre-COVID-19 cohort and a PCR-negative clinical cohort of patients presenting with COVID-19-like symptoms.

## Materials and methods

### Samples and cohorts

#### Serum samples from three different cohorts were used

1. PCR-positive clinical cohort: Samples from 29 patients, admitted to the hospital of LMU Munich with acute COVID-19 confirmed by positive PCR were collected over time from leftover material of samples submitted to our Institute for routine laboratory diagnostics. We collected serial samples from each patient (between 7 and 30 time points) covering a period of up to 64 days from the start of symptoms, adding up to a total of 443 samples (Fig 1). Samples were stored at −80°C as 250 µl aliquots in 2D barcoded biobanking vials (Thermo Scientific, Waltham, Massachusetts, USA) in the LMU LabMed Biobank. Clinical data of the PCR-positive clinical cohort (sex, age, date of symptom onset, date of first positive PCR, sepsis, immunosuppression, ARDS, death) were retrieved from electronic patient records. The patients were sampled at both regular wards and intensive care units. Anonymized analysis has been approved by the Ethics Committee of LMU Munich (reference number 20-552).
2. Pre-COVID-19 cohort: Samples from 238 healthy pre-COVID-19 subjects were collected from 04/2016 until 10/2019 as part of the Munich Study on Biomarker Reference Values (MyRef) for establishing age dependent reference values for laboratory tests. All samples originated from healthy individuals between 18 and 80 years without pre-existing conditions, pregnancy, lactation, smoking, excessive alcohol use or long-term medication (except oral contraceptives). The study has been approved by the Ethics Committee of LMU Munich (reference number 11/16), and written informed consent has been obtained from all participants.
3. PCR-negative clinical cohort: Samples from 257 patients, admitted to the hospital of LMU Munich with possible symptoms of SARS-CoV-2 but with a negative PCR result were collected from leftover material of samples submitted to our Institute for routine laboratory diagnostics. Samples were stored at −80°C as 250 µl aliquots in 2D barcoded biobanking vials (Thermo Scientific, Waltham, Massachusetts, USA) in the LMU LabMed Biobank. Demographic data were obtained from the electronic patient records. Anonymized analysis has been approved by the Ethics Committee of LMU Munich (reference number 20-552).

**Fig 1.**
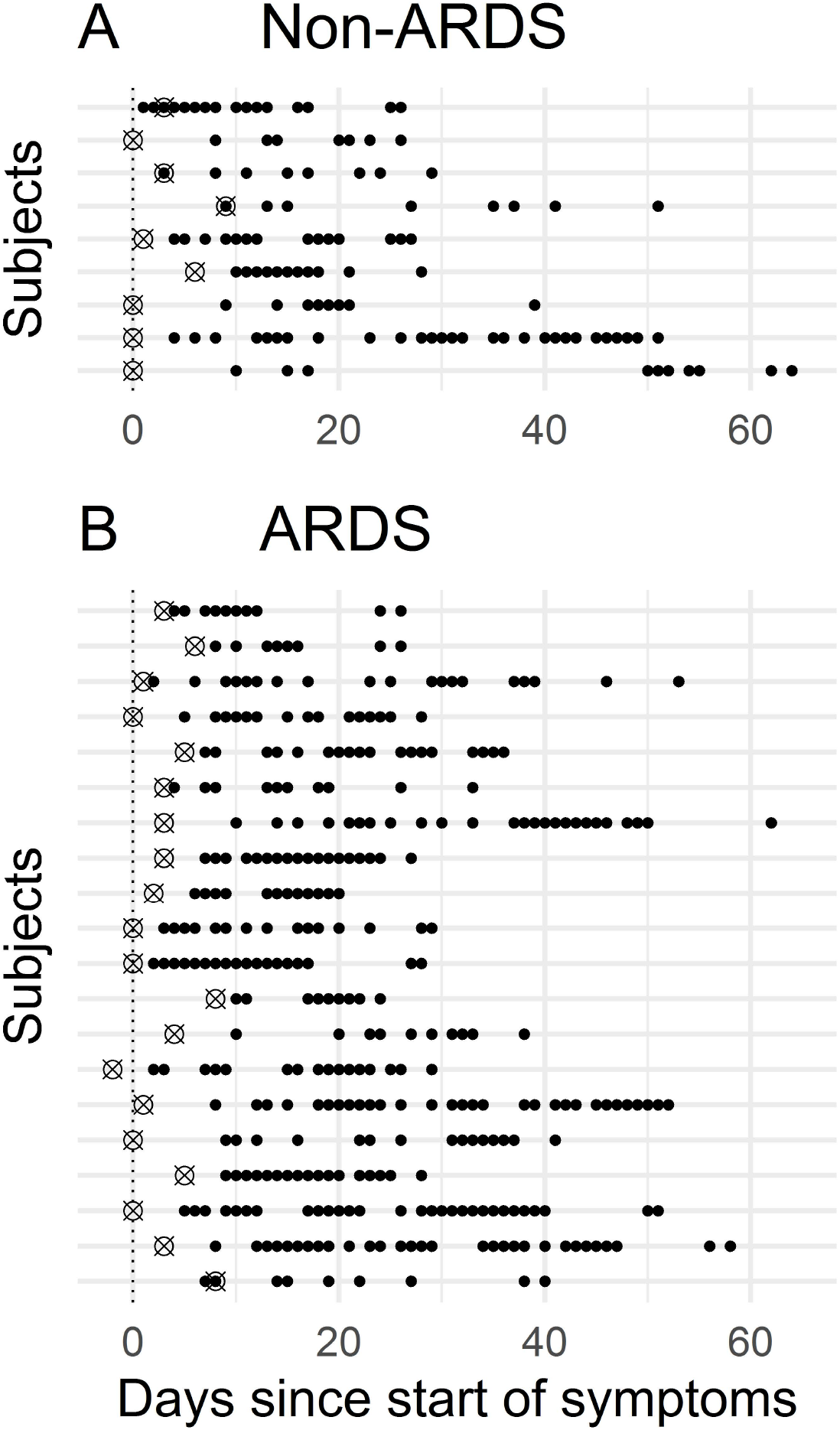
Time course of COVID-19 patient sampling. Day 0 represents symptom onset, crosses indicate the time of the first positive PCR result, dots indicate individual samples. (A) Non-ARDS patients. (B) ARDS patients.

### Serological assays

SARS-CoV-2 antibodies were analyzed using four commercially available immunoassays on analytical platforms, which are part of the operations for routine laboratory services provided by our Institute.

EUR S-IgA, EUR S-IgG and EUR N-IgG assays were semiquantitative enzyme-linked immunosorbent assays (ELISA) from Euroimmun (Euroimmun AG, Lübeck, Germany). EUR S-IgA and EUR S-IgG detect IgA and IgG against a recombinant S1 domain of the SARS-CoV-2 spike protein. EUR N-IgG detects IgG against a modified nucleocapsid protein. Assays were run on the fully automated ELISA processing platform Analyzer I (Euroimmun AG, Lübeck, Germany) according to the manufacturer’s instructions. Optical extinctions were normalized to an internal assay calibrator and reported as signal ratios between samples and calibrators. Signal ratios ≥ 1.1 were designated positive as suggested by the manufacturer. Values of the EUR S-IgA exceeding the upper limit were calculated as upper limit plus one. The Roche-Ab assay was a semiquantitative electrochemiluminiscence immunoassay from Roche Diagnostics (Roche Diagnostics International AG, Rotkreuz, Switzerland). It detected antibodies (including IgG) against a renatured chaperone nucleocapsid fusion protein. The assay was run on a cobas 8000 e 801 automated analyzer according to the manufacturer’s instructions. Results were reported as signal ratio between samples and cutoff calibrator. Signal ratios ≥ 1.0 were designated positive as suggested by the manufacturer. All samples were measured using all four immunoassays and expressed as qualitative result and as semiquantitative signal ratio. Individual values were considered to remain constant until the next measurement in the same individual.

### Statistics

Statistical analysis was performed with RStudio (R version 3.6.1) using the package ‘stats’. Patient subgroups in the PCR-positive clinical cohort were evaluated by their characteristics using the Fisher’s exact test or the Mann-Whitney-U test. Sensitivities were compared by points in time using the McNemar test for paired data. Sensitivities between different patient subgroups were determined using the Fishers exact test. Confidence intervals were calculated using an exact binomial test. Mean signal ratios between patient subgroups were compared by points in time using Welch’s t-test. Seroconversion times between patient subgroups were analyzed using the Mann-Whitney-U test. Specificities between different assays were compared using the McNemar test for paired data. Signal ratios were correlated using Spearman’s correlation coefficient. Age differences between SARS-CoV-2 antibody positive and negative patients in the negative cohort were analyzed using the Mann-Whitney-U test.

## Results

Patients’ characteristics are shown in Table 1. Patients were stratified according to diagnosis into groups without and with ARDS. The patients’ characteristics did not differ significantly between the two groups (Table 1).

**Table 1.**
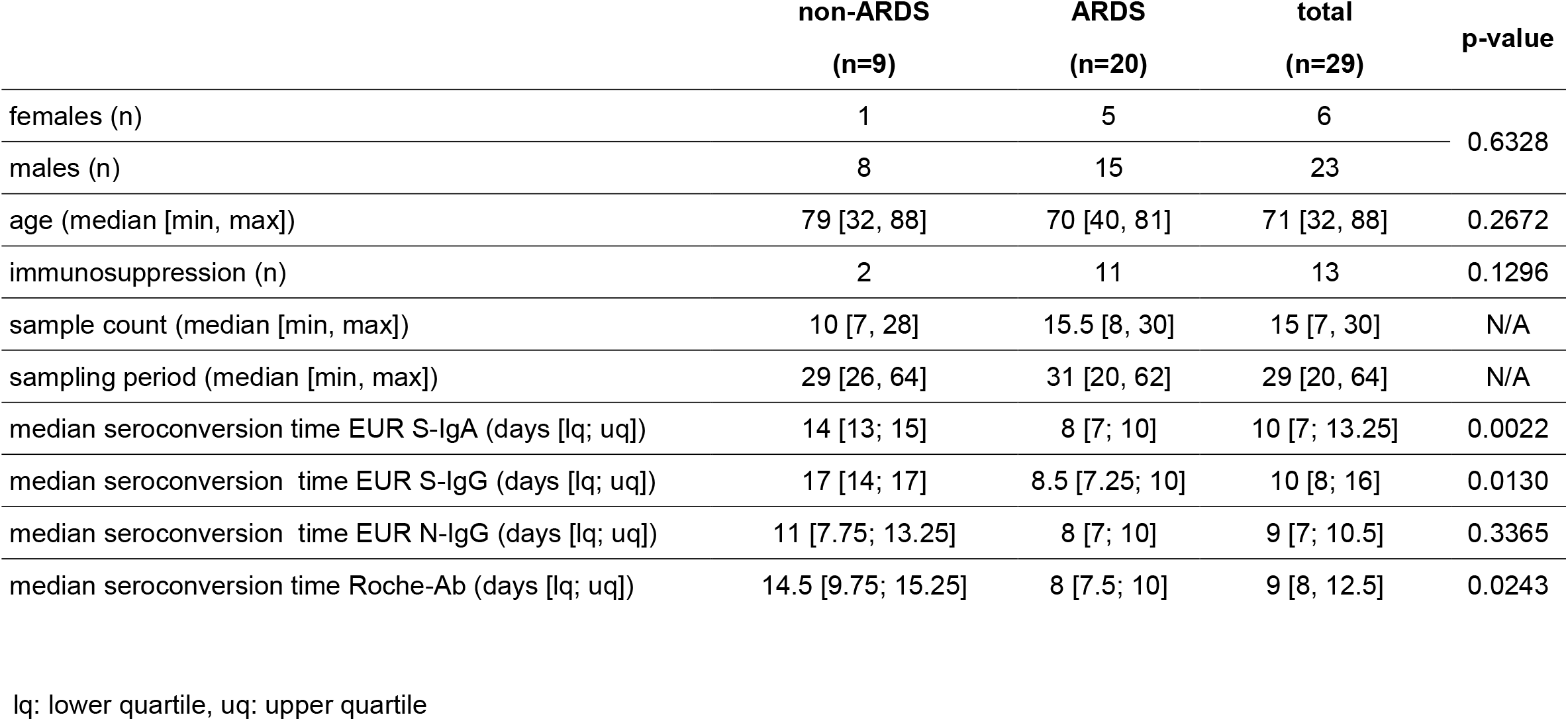
Characterization of the PCR-positive clinical cohort and stratification into non-ARDS and ARDS patients with respective median seroconversion times.

We first determined sensitivities of the four immunoassays categorized into positive and negative test results over time. Median seroconversion times for EUR S-IgA, EUR S-IgG, EUR N-IgG and Roche-Ab were comparable and are represented in Table 1. The dynamics of test sensitivities for all patients from day 5 to day 40 after symptom onset is shown in Fig 2A. Sensitivities for the different assays increased from between 0% and 25% at day 5 to between 90% and 97% at day 40 after symptom onset (Fig 2A). No significant difference between sensitivities of the four immunoassays was found, except for day 16 where EUR N-IgG was more sensitive than EUR S-IgG (p=0.041) (S1 Fig). Likewise, no significant differences between sensitivities of the four immunoassays were found when grouping time points into bins (S1 Table).

**Fig 2.**
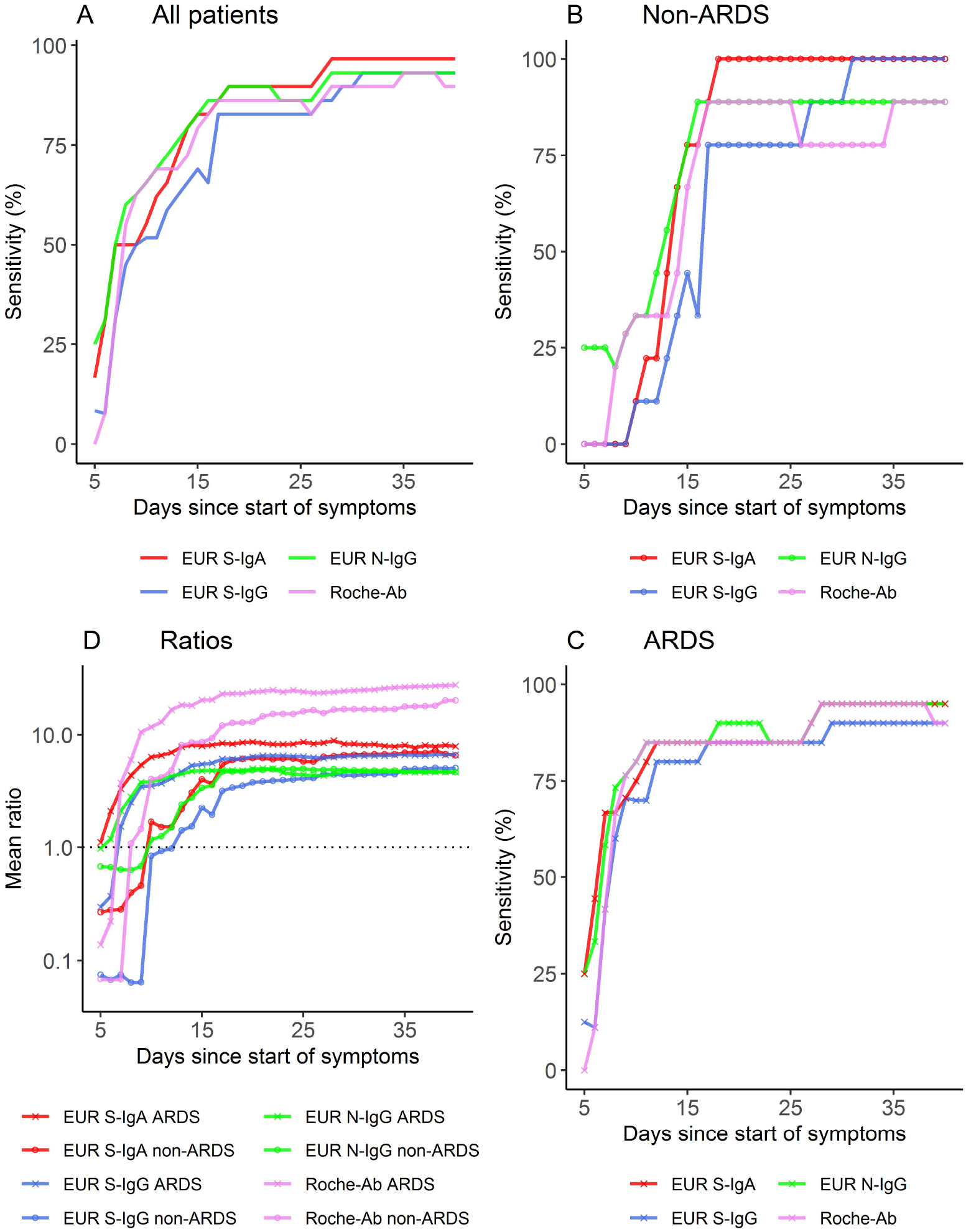
Immunoassay results over the course of time after symptom onset. Sensitivity in all patients. (B) Sensitivity in non-ARDS patients. (C) Sensitivity in ARDS patients. (D) Mean signal ratios of the four immunoassays in non-ARDS patients and ARDS patients.

We next examined the results of the four immunoassays in subgroups of non-ARDS and ARDS patients. Importantly, median seroconversion times were only 8-9 days in ARDS patients and 11-17 days in non-ARDS patients. The difference in seroconversion time was statistically significant between ARDS patients and non-ARDS patients for all immunoassays except EUR N-IgG (p=0.002 for EUR S-IgA, p=0.013 for EUR S-IgG, p=0.337 for EUR N-IgG, p=0.024 for Roche-Ab) (Table 1).

In addition, ARDS patients reached positivity at an earlier point in time than non-ARDS patients, as shown by significantly higher rates of positivity of the different tests between days 8 and 16 after symptom onset in the ARDS group (Figs 2B, 2C and 3). EUR S-IgG and EUR S-IgA discriminated between the two subgroups as early as day 8, whereas EUR N-IgG and Roche-Ab discriminated between the two subgroups starting on day 10 (Fig 3A). Mean signal ratios were significantly higher between days 7 to 19 for ARDS patients (Fig 3B).

**Fig 3.**
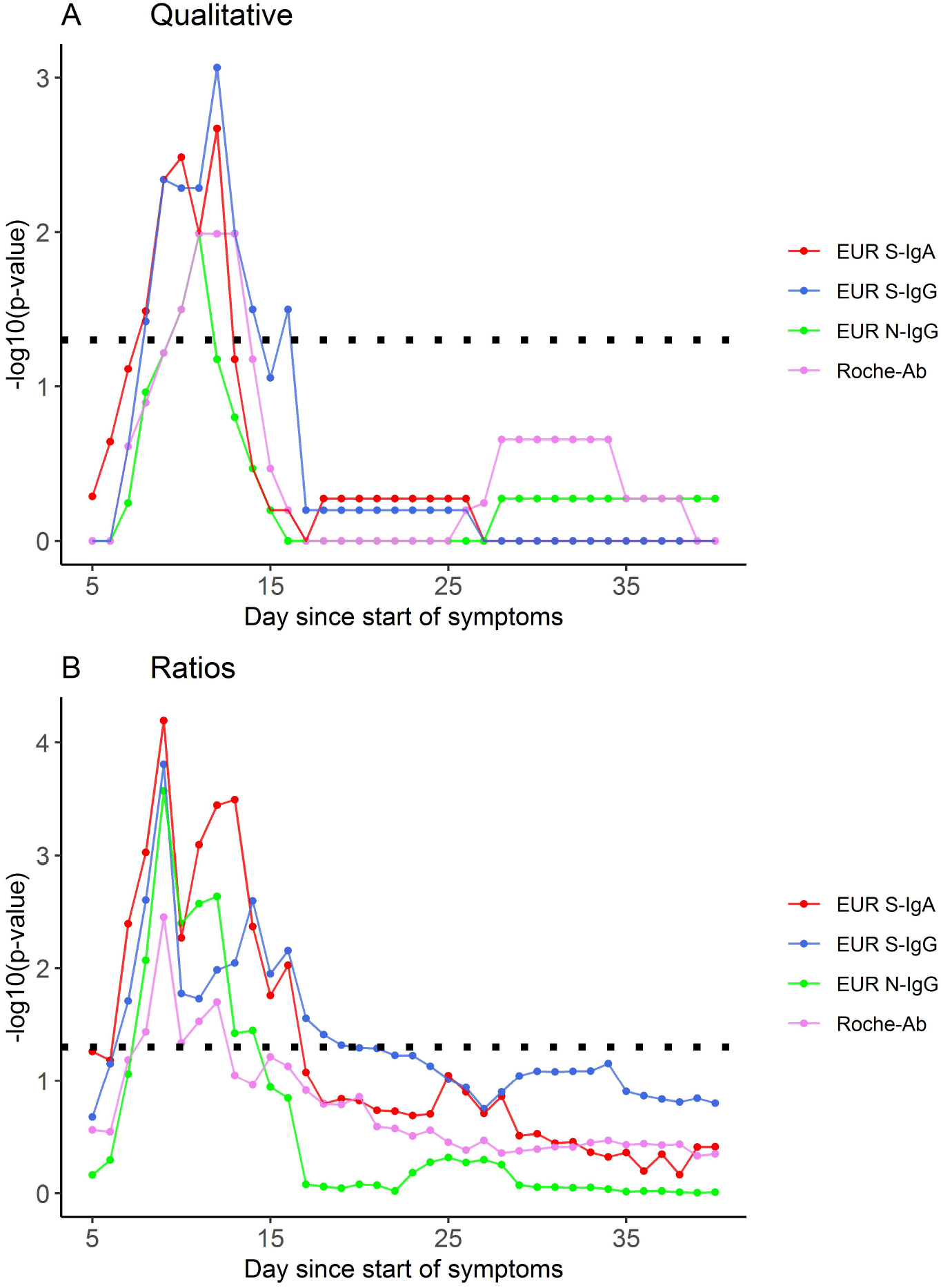
p-values of the differences between non-ARDS patients and ARDS patients for the four immunoassays at different time points. The dotted line represents a p-value of 0.05. (A) Differences in qualitative values. (B) Differences between mean signal ratios.

To further investigate this finding, we compared the mean signal ratios between the two subgroups over the course of time (Fig 2D). Significant differences in mean signal ratio were found as early as day 7 after symptom onset for EUR S-IgA and EUR S-IgG (p= 0.004, p=0.019) and day 8 for EUR N-IgG and Roche-Ab (p=0.008, p=0.037) (Fig 3B), corroborating our prior observations for the qualitative results. Additionally, we found significant differences of signal ratios between deceased and surviving patients for EUR S-IgG, EUR S-IgA and EUR N-IgG (days 12-35), and between septic and non-septic patients for Roche-Ab on day 28 (S2 Fig.). No differences were found between these groups for qualitative results. Pairwise comparison of signal ratios between the different immunoassays was performed and correlations are shown in S3 Fig. Distributions of signal ratios are shown in S4 Fig. The individual dynamics of signal ratios and clinical characterization for each patient are visualized in S5-S7 Figs. Individual dynamics of qualitative results and clinical characterization for each patient are visualized in S8-S10 Figs. One patient showed no antibody response at all. Another patient showed an antibody response only with the S-protein based immunoassays, whereas the Roche-Ab and EUR N-IgG immunoassays remained negative over the course of 64 days since onset of symptoms. One patient converted from positive to negative results only in the Roche-Ab immunoassay at day 39. Another immunosuppressed patient showed negative results only in the EUR S-IgG immunoassay over the period of 28 days since symptom onset.

We also determined the specificities of the four immunoassays (EUR S-IgA, EUR S-IgG, EUR N-IgG, Roche-Ab) using a pre-COVID-19 cohort of healthy individuals and in a PCR-negative clinical cohort of patients presenting with COVID-19-like symptoms (Table 2). Three subjects in the PCR-negative clinical cohort showed a positive result in all four immunoassays, suggesting that they had been previously exposed to SARS-CoV-2, and were therefore excluded from the following analysis. In the pre-COVID-19 cohort, EUR S-IgA consistently showed lower specificities than all other tests (p=0.00014, p=0.00014, p=0.00002). In the PCR-negative clinical cohort, EUR S-IgA showed lower specificities when compared to EUR S-IgG and Roche-Ab, but not when compared to EUR N-IgG (p=0.016, p=0.004, p=0.181) (Table 3). False positive results did not overlap between immunoassays except for one sample in the pre-COVID-19 cohort that showed a positive result in all three Euroimmun immunoassays, but not in the Roche immunoassay (S11 Fig).

**Table 2.** Antibody reactivity and specificity of the four immunoassays in two negative cohorts.

| assay | test result | pre-COVID19 cohort |  |  | PCR-negative clinical cohort |  |  | combined |
| --- | --- | --- | --- | --- | --- | --- | --- | --- |
|  |  | female<br>(n=146) | male<br>(n=92) | total<br>(n=238) | female<br>(n=115) | male<br>(n=142) | total<br>(n=257) |  |
| **EUR S-IgA** | neg. | 139 (95.2%) | 79 (85.9%) | 218 (91.6%) | 114 (99.1%) | 133 (93.7%) | 247 (96.1%) | 465 (93.9%) |
|  | pos. | 7 (4.8%) | 13 (14.1%) | 20 (8.4%) | 1 (0.9%) | 9 (6.3%) | 10 (3.9%) | 30 (6.1%) |
|  | Specificity (%) | 95.2 | 85.9 | 91.6 | 99.1 | 93.7 | 96.1 | 93.9 |
|  | [95% CI] | 90.4-98.1 | 77.0-92.3 | 87.3-94.8 | 95.3-100.0 | 88.3-97.1 | 93.0-98.1 | 91.5-95.9 |
| **EUR S-IgG** | neg. | 146 (100%) | 90 (97.8%) | 236 (99.2%) | 115 (100.0%) | 141 (99.3%) | 256 (99.6%) | 492 (99.4%) |
|  | pos. | 0 (0%) | 2 (2.2%) | 2 (0.8%) | 0 (0%) | 1 (0.7%) | 1 (0.4%) | 3 (0.6%) |
|  | Specificity (%) | 100.0 | 97.8 | 99.2 | 100.0 | 99.3 | 99.6 | 99.4 |
|  | [95% CI] | 97.5-100.0 | 92.4-99.7 | 97.0-99.9 | 96.8-100.0 | 96.1-100.0 | 97.9-100.0 | 98.2-99.9 |
| **EUR N-IgG** | neg. | 146 (100%) | 90 (97.8%) | 236 (99.2%) | 112 (97.4%) | 141 (99.3%) | 253 (98.4%) | 489 (98.8%) |
|  | pos. | 0 (0%) | 2 (2.2%) | 2 (0.8%) | 3 (2.6%) | 1 (0.7%) | 4 (1.6%) | 6 (1.2%) |
|  | Specificity (%) | 100.0 | 97.8 | 99.2 | 97.4 | 99.3 | 98.4 | 98.8 |
|  | [95% CI] | 97.5-100.0 | 92.4-99.7 | 97.0-99.9 | 92.6-99.5 | 96.1-100.0 | 96.1-99.6 | 97.4-99.6 |
| **Roche-Ab** | neg. | 146 (100%) | 92 (100%) | 238 (100%) | 115 (100.0%) | 142 (100.0%) | 257 (100.0%) | 495 (100.0%) |
|  | pos. | 0 (0%) | 0 (0%) | 0 (0%) | 0 (0.0%) | 0 (0.0%) | 0 (0.0%) | 0 (0.0%) |
|  | Specificity (%) | 100.0 | 100.0 | 100.0 | 100 | 100 | 100 | 100 |
|  | [95% CI] | 97.5-100.0 | 96.1-100.0 | 98.5-100.0 | 96.8-100.0 | 97.4-100.0 | 98.6-100.0 | 99.3-100.0 |

neg.: negative, pos.: positive, CI: confidence interval

**Table 3.** p-values of pairwise comparisons between specificities of immunoassays in the two sets of negative samples and both sets analyzed together (combined).

|  | pre-Covid-19 cohort | PCR-negative clinical cohort | combined |
| --- | --- | --- | --- |
| S-IgA : S-IgG | $1.4 \times 10^{-4}$ | $1.6 \times 10^{-2}$ | $3.0 \times 10^{-6}$ |
| S-IgA : N-IgG | $1.4 \times 10^{-4}$ | $1.8 \times 10^{-1}$ | $8.0 \times 10^{-5}$ |
| S-IgA : Roche-Ab | $2.1 \times 10^{-5}$ | $4.4 \times 10^{-3}$ | $1.2 \times 10^{-7}$ |
| N-IgG : S-IgG | 1.0 | $3.7 \times 10^{-1}$ | $4.5 \times 10^{-1}$ |
| S-IgG : Roche-Ab | $4.8 \times 10^{-1}$ | 1.0 | $2.5 \times 10^{-1}$ |
| N-IgG : Roche-Ab | $4.8 \times 10^{-1}$ | $1.3 \times 10^{-1}$ | $4.1 \times 10^{-2}$ |

The immunoassay with lower specificity is mentioned first

When performing pairwise comparisons between tests in the complete set of 495 negative samples, we found that EUR S-IgA consistently showed significantly lower specificities compared to the three other immunoassays (p ≤ 0.00008) (Table 3). In addition, EUR N-IgG also showed a significantly lower specificity than Roche-Ab (p=0.041). Furthermore, we found a significant age difference between false positive subjects and negative subjects in the EUR S-IgA and the EUR N-IgG immunoassay (p=0.029 and p=0.025, respectively) (S2 Table). Distributions of signal ratios are shown in S12 and S13 Figs.

## Discussion

Using serial serum sampling in patients hospitalized for COVID-19, we found that patients who developed ARDS in the course of disease had a substantially shorter seroconversion time (8–9 days) compared to patients who did not develop ARDS (11-17 days) (Table 1). This finding was consistent for IgA, IgG and total antibody responses. Earlier seropositivity in ARDS patients was confirmed by significantly higher rates of positivity of the different tests between days 8-16 after symptom onset (Figs 2B, 2C and 3). These findings were consistent for qualitative and semiquantitative analysis. Evidence for an earlier seroconversion in severe compared to mild cases has previously been reported by Yongchen et al. [24]. Similar to ours, this study used a serial sampling strategy, but included only a very limited number of samples and cases, providing no evidence for statistical significance of the findings. Another study analyzed IgM and total antibodies in single or serial samples from hospitalized patients and found that severe cases had significantly higher SARS-CoV-2 RBD-S1 antibody levels after day 6 from symptom onset [21]. While only few studies suggest earlier seroconversion in severe cases, a larger number reported an association of increased antibody levels with disease severity, intensive care unit status, and hospitalization [3, 4, 25] corroborating our findings for ARDS. In contrast, one study also reported a lack of association between antibody levels and disease severity [26], however, this study was only based on 15 PCR-positive cases with 2 to 6 serial measurements and might have been underpowered [26]. The National SARS-CoV-2 Serology Assay Evaluation Group could not find any evidence of a difference in sensitivity between immunoassays with regard to disease severity. However, the immunoassays investigated were different from our study, with the exception of the Roche-Ab immunoassay [27]. Median seroconversion times in our complete cohort of COVID-19 patients were 9-10 days since symptom onset for IgA, IgG and total antibodies, which is in line with other studies, reporting a seroconversion time of 7-14 days for IgM and IgG since symptom onset [3, 4, 6]. We found no differences of sensitivity in the early phase of infection suggesting that the detected antibody subtypes (IgA, IgG or total antibodies) seroconvert simultaneously. A simultaneous rise of all antibody subtypes [6] or an earlier rise of IgG in COVID-19 patients [5] has also been observed by others, and differs from infections with other agents, where IgM and IgA rise first and are markers of acute disease.

Furthermore, we found no significant differences in the sensitivity between the four immunoassays over the course of time (except for day 16). However, this question is an issue of current debate. Whereas Van Elslande et al. [28] report a faster seroconversion for N-protein based than for S-protein based SARS-CoV-2-antibody detection, a lack of a significant difference between S-based and N-based immunoassays regarding IgG and total antibodies was also found by others [29, 30].

When grouping the time points into bins, the EUR S-IgA, EUR N-IgG and Roche-Ab reached highest rates of sensitivity (92.9% – 96.4%) on days 20-29, the EUR S-IgG on days 30-39 (92.3%). EUR S-IgA sensitivities ranged from 88.0 % to 100.0% in other studies [13, 31-33], while EUR S-IgG ranged from 85.4% to 100% [32, 34]. Herroelen et al. [31] and Weidner et al. [35] found a sensitivity for EUR N-IgG of 90.5% and 88.9% respectively. Sensitivity for Roche-Ab ranged from 89.2% to 100% [36-38]. Therefore, peak levels of sensitivities found in our study were well within the range of others studies, except for EUR N-IgG, where we found a higher sensitivity. However, data on sensitivity of EUR N-IgG are still scarce. While we did not find any differences in the time binned sensitivities between the investigated immunoassays in our study, Meyer et al. found a significantly higher seropositivity for IgA (91.1%) than IgG (84.8%) 11-20 days after symptom onset with the EUR S-protein based ELISAs [13].

The analysis of individual dynamics of signal ratios and sensitivities showed one patient who developed no antibody response at all with all four tested immunoassays. This may be explained by immunosuppressive treatment of that patient. However, our study also included other patients with immunosuppressive therapy who clearly showed an antibody response. Therefore, the type of immunosuppressive therapy may be worth considering in further studies. Another patient showed an antibody response only with the S-protein based immunoassays, but not with the N-protein based EUR N-IgG and Roche-Ab (S8-S10 Figs). This finding is related to the controversial topic, whether antibodies against the N-or S-protein have higher sensitivity and rise earlier in the course of disease. From the analysis of individual antibody dynamics however, we cannot derive an answer to that question. It may be noted that some studies use the start of symptoms while others use positive PCR testing dates to monitor the occurrence of antibodies. We deemed the date of the first positive PCR as less reliable than the start of symptoms, because it is more dependent on external factors and therefore rather focused on the start of symptoms.

When analyzing the specificities in two COVID-19 negative cohorts, we found that the EUR S-IgA immunoassay consistently showed a lower specificity compared to the three other immunoassays. This finding is in line with other studies reporting a lower specificity of EUR S-IgA as well [2, 32, 33, 39-41]. Importantly, we found a significant difference in age distribution between the group of false positive subjects and true negative subjects for EUR S-IgA and EUR N-IgG results. The mean age of false positives was higher than the mean age of true negatives for EUR N-IgG, and the other way around for EUR S-IgA (S2 Table). It has been speculated that older populations might have higher cross-reactivities due to a longer history of interaction with other types of coronaviruses [12]. Accordingly, Gorse et al. reported that over 90% of adults over age 50 present antibodies to four common circulating coronaviruses [42]. This might explain our result regarding age differences of false positives for EUR N-IgG, but not for EUR S-IgA.

With regard to specificity, the EUR S-IgG and EUR N-IgG showed similar results in our study. Specificities for the EUR S-IgG and EUR N-IgG immunoassays (99.4% and 98.8%, respectively) were in line with a number of studies [13, 31, 43, 44], while others also reported lower specificities for EUR S-IgG (91.9% −96.2%) [39, 45, 46]. In our study, the Roche-Ab immunoassay, which demonstrated 100% specificity, had a significantly higher specificity than EUR S-IgA and EUR N-IgG in the total negative cohort. Similarly, others reported 100 % specificity of the Roche-Ab immunoassay as well [38, 47, 48], whereas Ekelund et al. found a specificity of 98 % [37]. Despite our relatively high number of samples in the PCR positive cohort, the absolute number of individual subjects is limited. Therefore, it will be interesting to see whether these results can be replicated in independent, prospective cohorts.

In conclusion, the specificities of the four SARS-CoV-2 immunoassays investigated in our study were higher for Roche-Ab and EUR S-IgG compared to EUR S-IgA and EUR N-IgG. In contrast, the sensitivities were comparable. Serial sampling revealed an early rise in SARS-CoV-2 IgA, IgG and total antibodies as a potential indicator of ARDS in COVID-19 patients. This finding suggests that SARS-CoV-2 antibodies may serve as biomarkers for early detection of ARDS, as a complication of COVID-19, and warrants replication in future studies.

## Data Availability

Data are available on request.

## Acknowledgments

We thank Britta Pauli and Babett Rannefeld for their expert technical assistance.

## Supporting information

**S1 Fig. p-values for differences in qualitative test results for the four immunoassays in the PCR-positive clinical cohort**. The dotted line represents a p-value of 0.05.

**S1 Table. Sensitivities (with 95% confidence interval) of the different immunoassays grouped into time bins**.

**S2 Fig. p-values for differences between patients by death and sepsis for the four immunoassays at different time points**. The dotted line represents a p-value of 0.05. (A) Differences in qualitative values for deceased and surviving patients.

(B) Differences between mean signal ratios for deceased and surviving patients.

(C) Differences in qualitative values for septic and non-septic patients.

(D) Differences between mean signal ratios for septic and non-septic patients.

**S3 Fig. Pairwise comparison of signal ratios between the different immunoassays in the PCR-positive clinical cohort**. Spearman correlation coefficient (R) and p-values are shown. The dotted lines represent the cutoff values for a positive test result.

**S4 Fig. Distributions of signal ratios for the four different immunoassays in the PCR-positive clinical cohort**. The dotted lines represent the cutoff values for a positive test result. **(A)** EUR S-IgA. **(B)** EUR S-IgG. **(C)** EUR N-IgG. **(D)** Roche-Ab.

**S5 Fig. Individual results in the PCR-positive clinical cohort for the four different immunoassays in the non-ARDS group**.

**S6 Fig. Individual results in the PCR-positive clinical cohort for the four different immunoassays in the ARDS group (first set)**.

**S7 Fig. Individual results in the PCR-positive clinical cohort for the four different immunoassays in the ARDS group (second set)**.

**S8 Fig. Individual qualitative results in the PCR-positive clinical cohort for the four different immunoassays in the non-ARDS group**.

**S9 Fig. Individual qualitative results in the PCR-positive clinical cohort for the four different immunoassays in the ARDS group (first set)**.

**S10 Fig. Individual qualitative results in the PCR-positive clinical cohort for the four different immunoassays in the ARDS group (second set)**.

**S11 Fig. Overlap of positive results between immunoassays in the two negative cohorts**. (A) Pre-COVID-19 cohort. (B) PCR-negative clinical cohort. (This plot was generated using the UpSetR R package)

**S2 Table. Median age for true negative and false positive subjects in the negative cohorts**

**S12 Fig. Distributions of signal ratios for the four different immunoassays in the pre-COVID-19 cohort**. The dotted lines represent the cutoff values for a positive test result. (A) EUR S-IgA. (B) EUR S-IgG. (C) EUR N-IgG. (D) Roche-Ab.

**S13 Fig. Distributions of signal ratios for the four different immunoassays in the PCR-negative clinical cohort**. The dotted lines represent the cutoff values for a positive test result. (A) EUR S-IgA. (B) EUR S-IgG. (C) EUR N-IgG. (D) Roche-Ab.

## References

1. Centers for Disease Control and Prevention, Coronavirus Disease 2019 (COVID-19), Overview of Testing for SARS-CoV-2 [accessed on 2020 Oct 15]. Available from: https://www.cdc.gov/coronavirus/2019-ncov/hcp/testing-overview.html.

2. Okba NM, Muller MA, Li W, Wang C, GeurtsvanKessel CH, Corman VM, et al. Severe acute respiratory syndrome coronavirus 2-specific antibody responses in coronavirus disease patients. Emerg Infect Dis 2020;26:1478–88.

3. Zhao J, Yuan Q, Wang H, Liu W, Liao X, Su Y, et al. Antibody responses to SARS-CoV-2 in patients of novel coronavirus disease 2019. Clin Infect Dis 2020 Mar 28. doi:10.1093/cid/ciaa344.

4. Tan W, Lu Y, Zhang J, Wang J, Dan Y, Tan Z, et al. Viral kinetics and antibody responses in patients with COVID-19. medRxiv [Preprint]. 2020 [posted 2020 Mar 26]. Available from: https://doi.org/10.1101/2020.03.24.20042382.

5. To KK, Tsang OT, Leung WS, Tam AR, Wu TC, Lung DC, et al. Temporal profiles of viral load in posterior oropharyngeal saliva samples and serum antibody responses during infection by SARS-CoV-2: an observational cohort study. Lancet Infect Dis 2020;20:565–74.

6. Long QX, Liu BZ, Deng HJ, Wu GC, Deng K, Chen YK, et al. Antibody responses to SARS-CoV-2 in patients with COVID-19. Nat Med 2020;26:845–848.

7. Cervia C, Nilsson J, Zurbuchen Y, Valaperti A, Schreiner J, Wolfensberger A, et al. Systemic and mucosal antibody responses specific to SARS-CoV-2 during mild versus severe COVID-19. Allergy Clin Immunol 2020;20:S0091-6749(20)31623-7.

8. Long QX, Tang XJ, Shi QL, Li Q, Deng HJ, Yuan J, et al. Clinical and immunological assessment of asymptomatic SARS-CoV-2 infections. Nat Med 2020;26:1200–4.

9. Sethuraman N, Jeremiah SS, Ryo A. Interpreting diagnostic tests for SARS-CoV-2. JAMA 2020;323(22):2249–2251.

10. Deeks JJ, Dinnes J, Takwoingi Y, Davenport C, Spijker R, Taylor-Phillips S, et al. Antibody tests for identification of current and past infection with SARS-CoV-2. Cochrane Database Syst Rev 2020;6:CD013652.

11. Theel ES, Slev P, Wheeler S, Couturier MR, Wong SJ, Kadkhoda K. The role of antibody testing for SARS-CoV-2: is there one? J Clin Microbiol 2020;58:e00797–20.

12. Farnsworth CW, Anderson NW. SARS-CoV-2 Serology: Much Hype, Little Data. Clin Chem 2020;66:875–7.

13. Meyer B, Torriani G, Yerly S, Mazza L, Calame A, Arm-Vernez I, et al. Validation of a commercially available SARS-CoV-2 serological immunoassay. Clin Microbiol Infect 2020;26:1386–1394.

14. Diamandis P, Prassas I, Diamandis EP. Antibody tests for COVID-19: drawing attention to the importance of analytical specificity. Clin Chem Lab Med 2020;58:1144–45.

15. Bohn MK, Lippi G, Horvath A, Sethi S, Koch D, Ferrari M, et al. Molecular, serological, and biochemical diagnosis and monitoring of COVID-19: IFCC taskforce evaluation of the latest evidence. Clin Chem Lab Med 2020;58:1037–52.

16. Shi J, Han D, Zhang R, Li J, Zhang R. Molecular and serological assays for SARS-CoV-2: insights from genome and clinical characteristics. Clin Chem 2020;66:1030–1046.

17. Zhou P, Yang XL, Wang XG, Hu B, Zhang L, Zhang W, et al. A pneumonia outbreak associated with a new coronavirus of probable bat origin. Nature 2020;579:270–73.

18. Lu R, Zhao X, Li J, Niu P, Yang B, Wu H, et al. Genomic characterisation and epidemiology of 2019 novel coronavirus: implications for virus origins and receptor binding. Lancet 2020;395(10224):565–74.

19. Guo L, Ren L, Yang S, Xiao M, Chang D, Yang F, et al. Profiling early humoral response to diagnose Novel Coronavirus Disease (COVID-19). Clin Infect Dis 2020;71:778–85.

20. Yang X, Yu Y, Xu J, Shu H, Xia J, Liu H, et al. Clinical course and outcomes of critically ill patients with SARS-CoV-2 pneumonia in Wuhan, China: a single-centered, retrospective, observational study. Lancet Respir Med 2020;8:475–81.

21. Liu ZL, Liu Y, Wan LG, Xiang TX, L. AP, Liu P, et al. Antibody profiles in mild and severe cases of COVID-19. Clin Chem 2020;66:1102–1104.

22. Robbiani DF, Gaebler C, Muecksch F, Lorenzi JCC, Wang Z, Cho A, et al. Convergent antibody responses to SARS-CoV-2 in convalescent individuals. Nature 2020;584(7821):437–42.

23. Chen X, Pan Z, Yue S, Yu F, Zhang J, Yang Y, et al. Disease severity dictates SARS-CoV-2-specific neutralizing antibody responses in COVID-19. Signal Transduct Target Ther 2020(1):180.

24. Yongchen Z, Shen H, Wang X, Shi X, Li Y, Yan J, et al. Different longitudinal patterns of nucleic acid and serology testing results based on disease severity of COVID-19 patients. Emerg Microbes Infect 2020;9:833–36.

25. Bonelli F, Sarasini A, Zierold C, Calleri M, Bonetti A, Vismara C, et al. Clinical and analytical performance of an automated serological test that identifies S1/S2 neutralizing IgG in COVID-19 patients semiquantitatively. J Clin Microbiol 2020;58(9):e01224–20.

26. Phipps WS, SoRelle JA, Li QZ, Mahimainathan L, Araj E, Markantonis J, et al. SARS-CoV-2 Antibody Responses Do Not Predict COVID-19 Disease Severity. Am J Clin Pathol 2020;154:459–65.

27. National S-C-SAEG. Performance characteristics of five immunoassays for SARS-CoV-2: a head-to-head benchmark comparison. Lancet Infect Dis 2020;20:1390–400.

28. Van Elslande J, Decru B, Jonckheere S, Van Wijngaerden E, Houben E, Vandecandelaere P, et al. Antibody response against SARS-CoV-2 spike protein and nucleoprotein evaluated by four automated immunoassays and three ELISAs. Clin Microbiol Infect 2020;26(11):1557.e1-1557.e7.2020.

29. Tang MS, Hock KG, Logsdon NM, Hayes JE, Gronowski AM, Anderson NW, et al. Clinical performance of the Roche SARS-CoV-2 serologic assay. Clin Chem 2020;66:1107–1109.

30. Liu W, Liu L, Kou G, Zheng Y, Ding Y, Ni W, et al. Evaluation of Nucleocapsid and Spike Protein-based ELISAs for detecting antibodies against SARS-CoV-2. J Clin Microbiol 2020;58(6):e00461–20.

31. Herroelen PH, Martens GA, De Smet D, Swaerts K, Decavele AS. Humoral Immune Response to SARS-CoV-2. Am J Clin Pathol 2020 Aug 18. doi:10.1093/ajcp/aqaa140.

32. Nicol T, Lefeuvre C, Serri O, Pivert A, Joubaud F, Dubee V, et al. Assessment of SARS-CoV-2 serological tests for the diagnosis of COVID-19 through the evaluation of three immunoassays: Two automated immunoassays (Euroimmun and Abbott) and one rapid lateral flow immunoassay (NG Biotech). J Clin Virol 2020;129:104511.

33. Charlton CL, Kanji JN, Johal K, Bailey A, Plitt SS, MacDonald C, et al. Evaluation of six commercial mid to high volume antibody and six point of care lateral flow assays for detection of SARS-CoV-2 antibodies. J Clin Microbiol 2020;58(10):e01361–20.

34. Tang MS, Hock KG, Logsdon NM, Hayes JE, Gronowski AM, Anderson NW, et al. Clinical performance of two SARS-CoV-2 serologic assays. Clin Chem 2020;66:1055–1062.

35. Weidner L, Gansdorfer S, Unterweger S, Weseslindtner L, Drexler C, Farcet M, et al. Quantification of SARS-CoV-2 antibodies with eight commercially available immunoassays. J Clin Virol 2020;129:104540.

36. Perkmann T, Perkmann-Nagele N, Breyer MK, Breyer-Kohansal R, Burghuber OC, Hartl S, et al. Side by side comparison of three fully automated SARS-CoV-2 antibody assays with a focus on specificity. Clin Chem 2020;66:1405–1413.

37. Ekelund O, Ekblom K, Somajo S, Pattison-Granberg J, Olsson K, Petersson A. High-throughput immunoassays for SARS-CoV-2, considerable differences in performance when comparing three methods. medRxiv [Preprint]. 2020 [posted 2020 Sep 17]. Available from: https://doi.org/10.1101/2020.05.22.20106294.

38. Egger M, Bundschuh C, Wiesinger K, Gabriel C, Clodi M, Mueller T, et al. Comparison of the Elecsys(R) Anti-SARS-CoV-2 immunoassay with the EDI enzyme linked immunosorbent assays for the detection of SARS-CoV-2 antibodies in human plasma. Clin Chim Acta 2020;509:18–21.

39. Jaaskelainen AJ, Kekalainen E, Kallio-Kokko H, Mannonen L, Kortela E, Vapalahti O, et al. Evaluation of commercial and automated SARS-CoV-2 IgG and IgA ELISAs using coronavirus disease (COVID-19) patient samples. Euro Surveill 2020;25:2000603.

40. Montesinos I, Gruson D, Kabamba B, Dahma H, Van den Wijngaert S, Reza S, et al. Evaluation of two automated and three rapid lateral flow immunoassays for the detection of anti-SARS-CoV-2 antibodies. J Clin Virol 2020;128:104413.

41. Jaaskelainen AJ, Kuivanen S, Kekalainen E, Ahava MJ, Loginov R, Kallio-Kokko H, et al. Performance of six SARS-CoV-2 immunoassays in comparison with microneutralisation. J Clin Virol 2020;129:104512.

42. Gorse GJ, Patel GB, Vitale JN, O’Connor TZ. Prevalence of antibodies to four human coronaviruses is lower in nasal secretions than in serum. Clin Vaccine Immunol 2010;17:1875–80.

43. Theel ES, Harring J, Hilgart H, Granger D. Performance characteristics of four high-throughput immunoassays for detection of IgG antibodies against SARS-CoV-2. J Clin Microbiol 2020;58:e01243–20.

44. Kohmer N, Westhaus S, Ruhl C, Ciesek S, Rabenau HF. Brief clinical evaluation of six high-throughput SARS-CoV-2 IgG antibody assays. J Clin Virol 2020;129:104480.

45. Van Elslande J, Houben E, Depypere M, Brackenier A, Desmet S, Andre E, et al. Diagnostic performance of seven rapid IgG/IgM antibody tests and the Euroimmun IgA/IgG ELISA in COVID-19 patients. Clin Microbiol Infect 2020;26:1082–87.

46. Kruttgen A, Cornelissen CG, Dreher M, Hornef M, Imohl M, Kleines M. Comparison of four new commercial serologic assays for determination of SARS-CoV-2 IgG. J Clin Virol 2020;128:104394.

47. Harb R, Remaley AT, Sacks DB. Evaluation of Three Commercial Automated Assays for the Detection of Anti-SARS-CoV-2 Antibodies. Clin Chem 2020;66:1351–3.

48. Favresse J, Eucher C, Elsen M, Tre-Hardy M, Dogne JM, Douxfils J. Clinical Performance of the Elecsys Electrochemiluminescent Immunoassay for the Detection of SARS-CoV-2 Total Antibodies. Clin Chem 2020;66:1104–6.

