## Supplementary material for "SARS-CoV-2 antibody immunoassays in serial samples reveal earlier seroconversion in acutely ill COVID-19 patients developing ARDS": S1 Fig

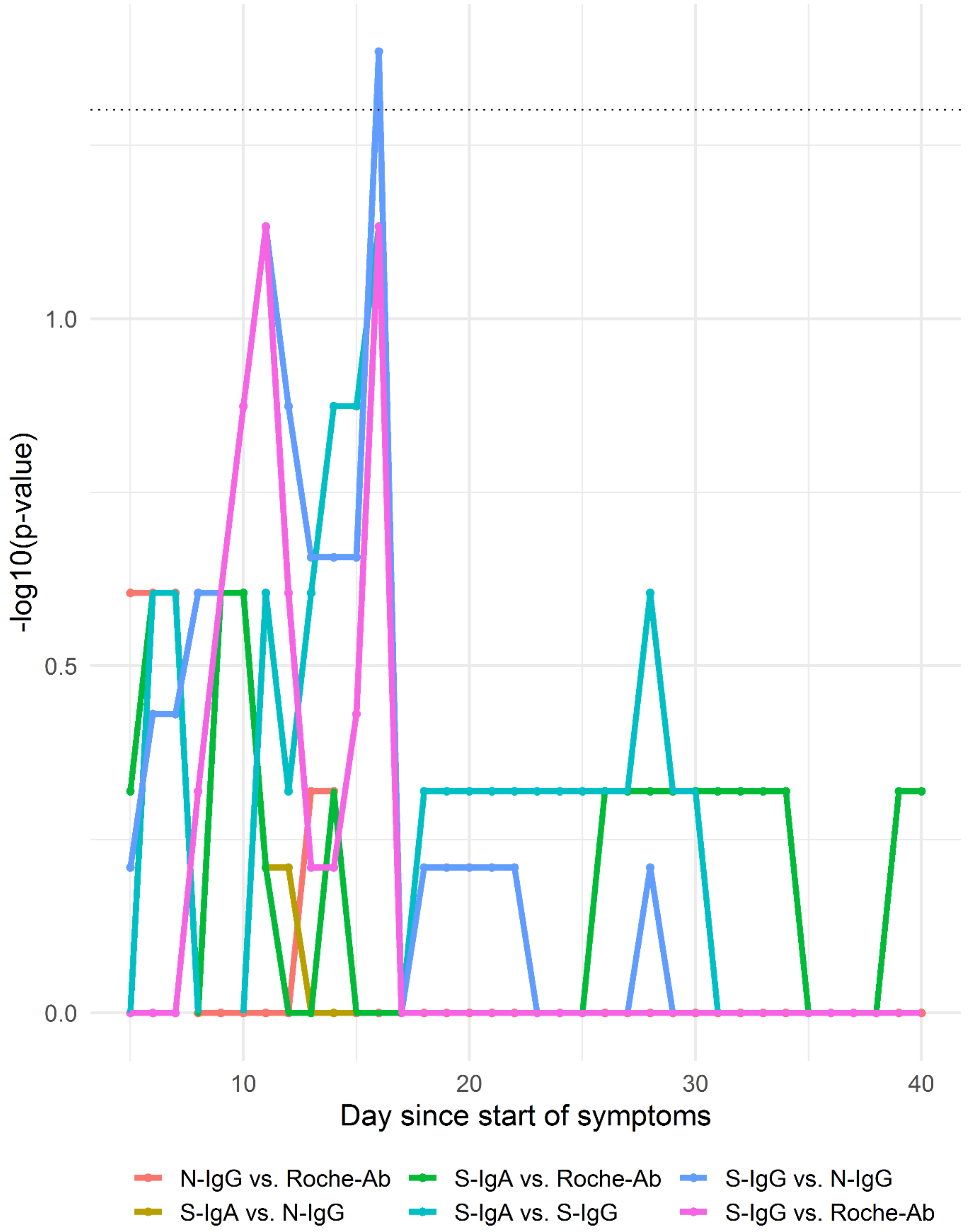


**S1 Fig. p-values for differences in qualitative test results for the four immunoassays in the PCR-positive clinical cohort.** The dotted line represents a p-value of 0.05.
