## Supplementary material for "SARS-CoV-2 antibody immunoassays in serial samples reveal earlier seroconversion in acutely ill COVID-19 patients developing ARDS": S1 Table

**S1 Table. Sensitivities (with 95% confidence interval) of the different immunoassays grouped into time bins.**

| time period | n | EUR S-IgA | EUR S-IgG | EUR N-IgG | Roche-Ab |
|---|---|---|---|---|---|
| <5 | 10 | 10.0 (0.3% to 44.5%) | 10.0 (0.3% to 44.5%) | 20.0 (2.5% to 55.6%) | 0.0 (0.0% to 30.8%) |
| 5-9 | 24 | 50.0 (29.1% to 70.9%) | 50.0 (29.1% to 70.9%) | 62.5 (40.6% to 81.2%) | 62.5 (40.6% to 81.2%) |
| 10-14 | 28 | 78.6 (59.0% to 91.7%) | 64.3 (44.1% to 81.4%) | 78.6 (59.0% to 91.7%) | 71.4 (51.3% to 86.8%) |
| 15-19 | 26 | 88.5 (69.8% to 97.6%) | 80.8 (60.6% to 93.4%) | 88.5 (69.8% to 97.6%) | 84.6 (65.1% to 95.6%) |
| 20-29 | 28 | 96.4 (81.7% to 99.9%) | 89.3 (71.8% to 97.7%) | 96.4 (81.7% to 99.9%) | 92.9 (76.5% to 99.1%) |
| 30-39 | 13 | 92.3 (64.0% to 99.8%) | 92.3 (64.0% to 99.8%) | 92.3 (64.0% to 99.8%) | 92.3 (64.0% to 99.8%) |
| 40-49 | 9 | 88.9 (51.8% to 99.7%) | 88.9 (51.8% to 99.7%) | 88.9 (51.8% to 99.7%) | 77.8 (40.0% to 97.2%) |
| 50-64 | 8 | 87.5 (47.3% to 99.7%) | 87.5 (47.3% to 99.7%) | 75.0 (34.9% to 96.8%) | 62.5 (24.5% to 91.5%) |
