## Supplementary material for "SARS-CoV-2 antibody immunoassays in serial samples reveal earlier seroconversion in acutely ill COVID-19 patients developing ARDS": S2 Fig

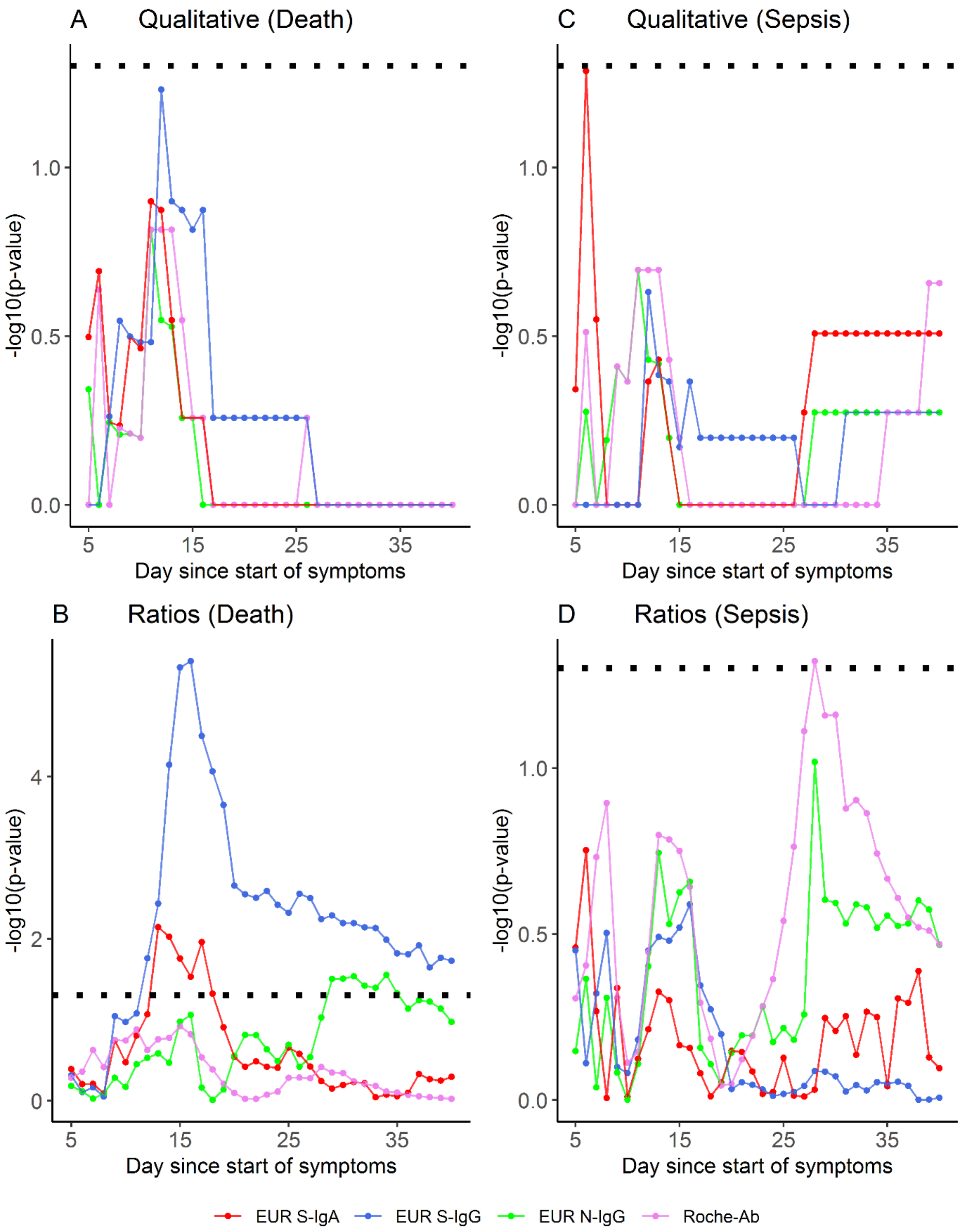


**S2 Fig. p-values for differences between patients by death and sepsis for the four immunoassays at different time points.** The dotted line represents a p-value of 0.05. (A) Differences in qualitative values for deceased and surviving patients. (B) Differences between mean signal ratios for deceased and surviving patients. (C) Differences in qualitative values for septic and non-septic patients. (D) Differences between mean signal ratios for septic and non-septic patients.
