## Supplementary material for "SARS-CoV-2 antibody immunoassays in serial samples reveal earlier seroconversion in acutely ill COVID-19 patients developing ARDS": S3 Fig

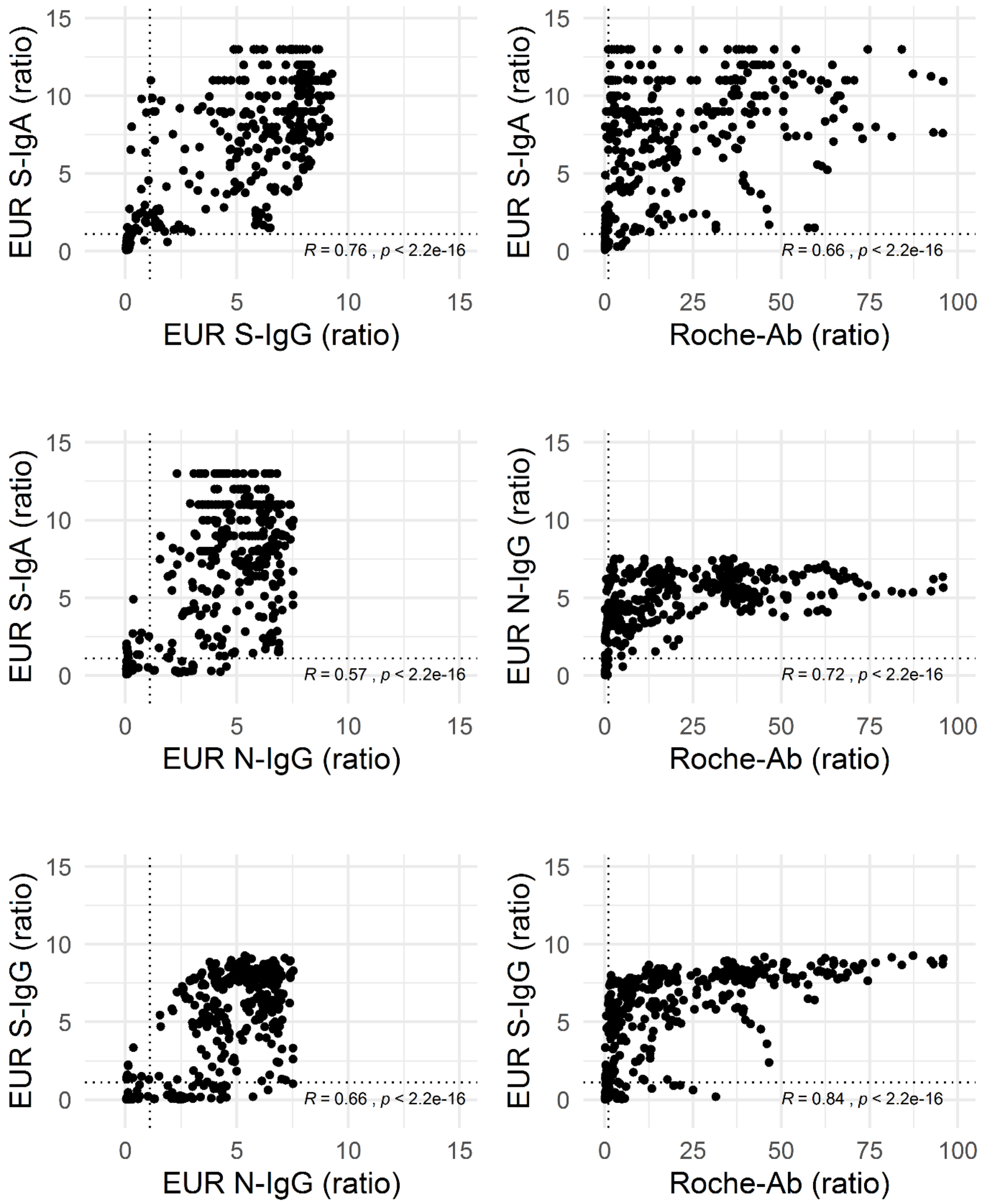


**S3 Fig. Pairwise comparison of signal ratios between the different immunoassays in the PCR-positive clinical cohort.** Spearman correlation coefficient (R) and p-values are shown. The dotted lines represent the cutoff values for a positive test result.
