## Supplementary material for "SARS-CoV-2 antibody immunoassays in serial samples reveal earlier seroconversion in acutely ill COVID-19 patients developing ARDS": S6 Fig

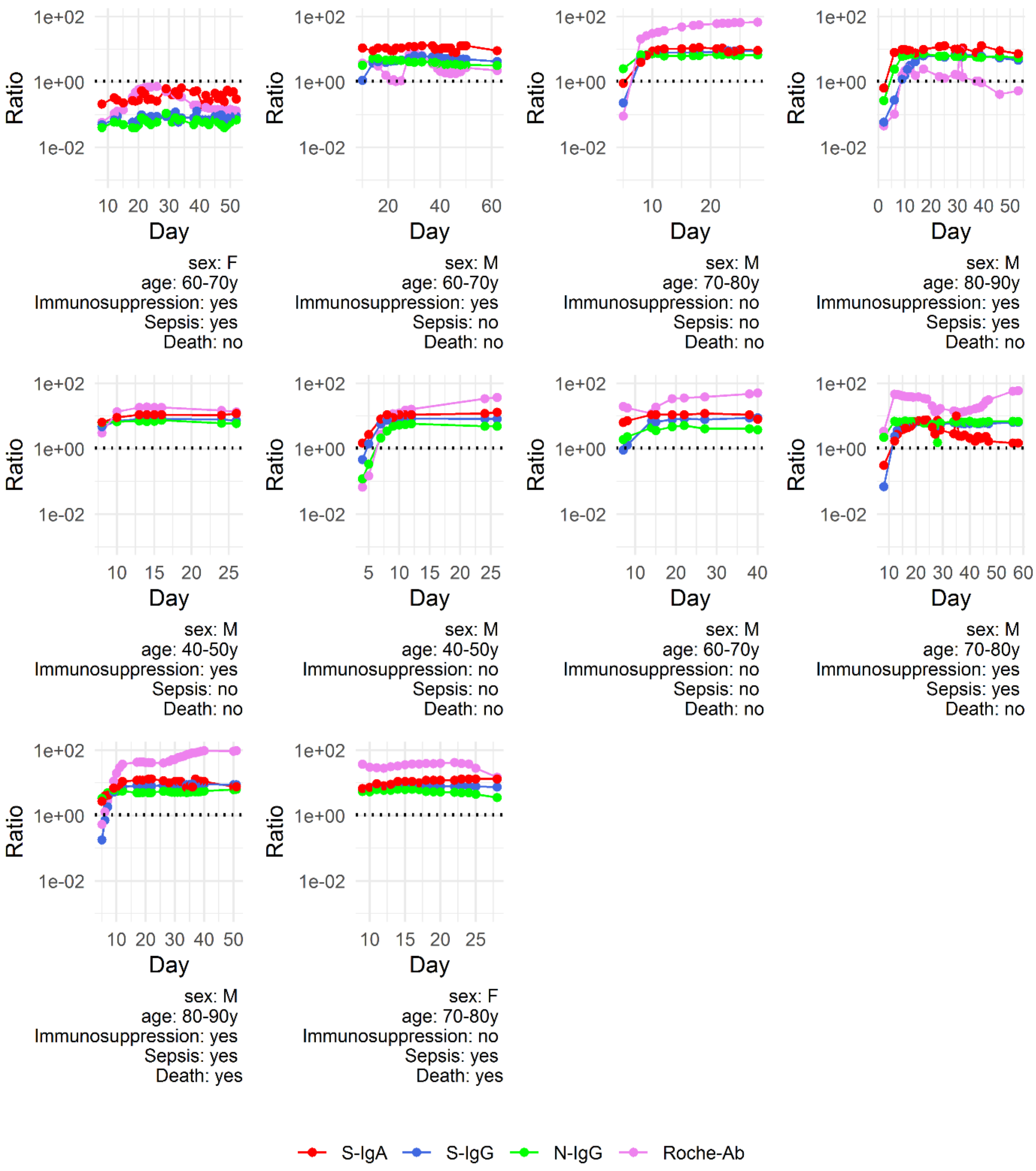


**S6 Fig. Individual results in the PCR-positive clinical cohort for the four different immunoassays in the ARDS group (first set).**
