## Supplementary material for "SARS-CoV-2 antibody immunoassays in serial samples reveal earlier seroconversion in acutely ill COVID-19 patients developing ARDS": S7 Fig

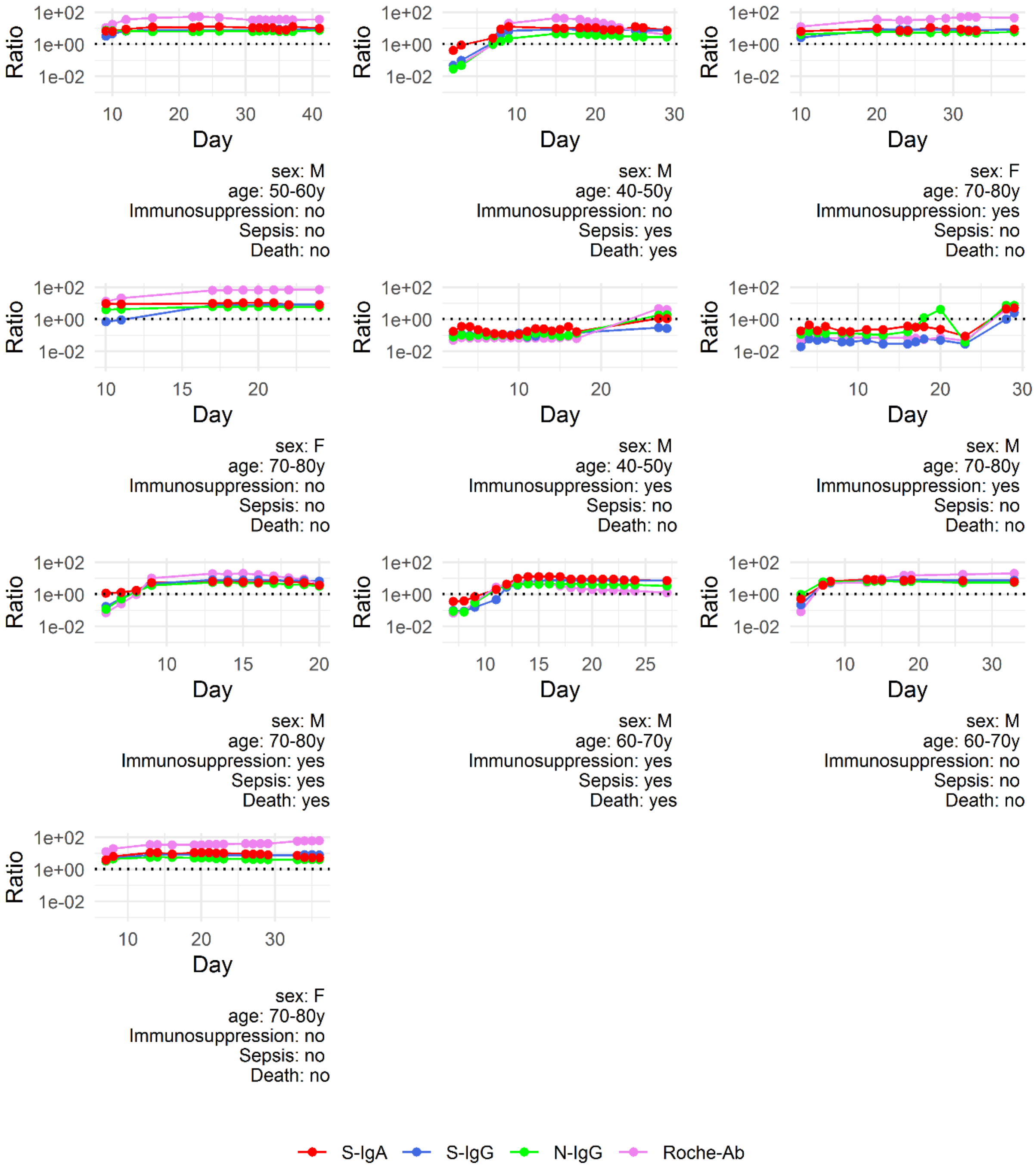


**S7 Fig. Individual results in the PCR-positive clinical cohort for the four different immunoassays in the ARDS group (second set).**
