## Supplementary material for "SARS-CoV-2 antibody immunoassays in serial samples reveal earlier seroconversion in acutely ill COVID-19 patients developing ARDS": S8 Fig

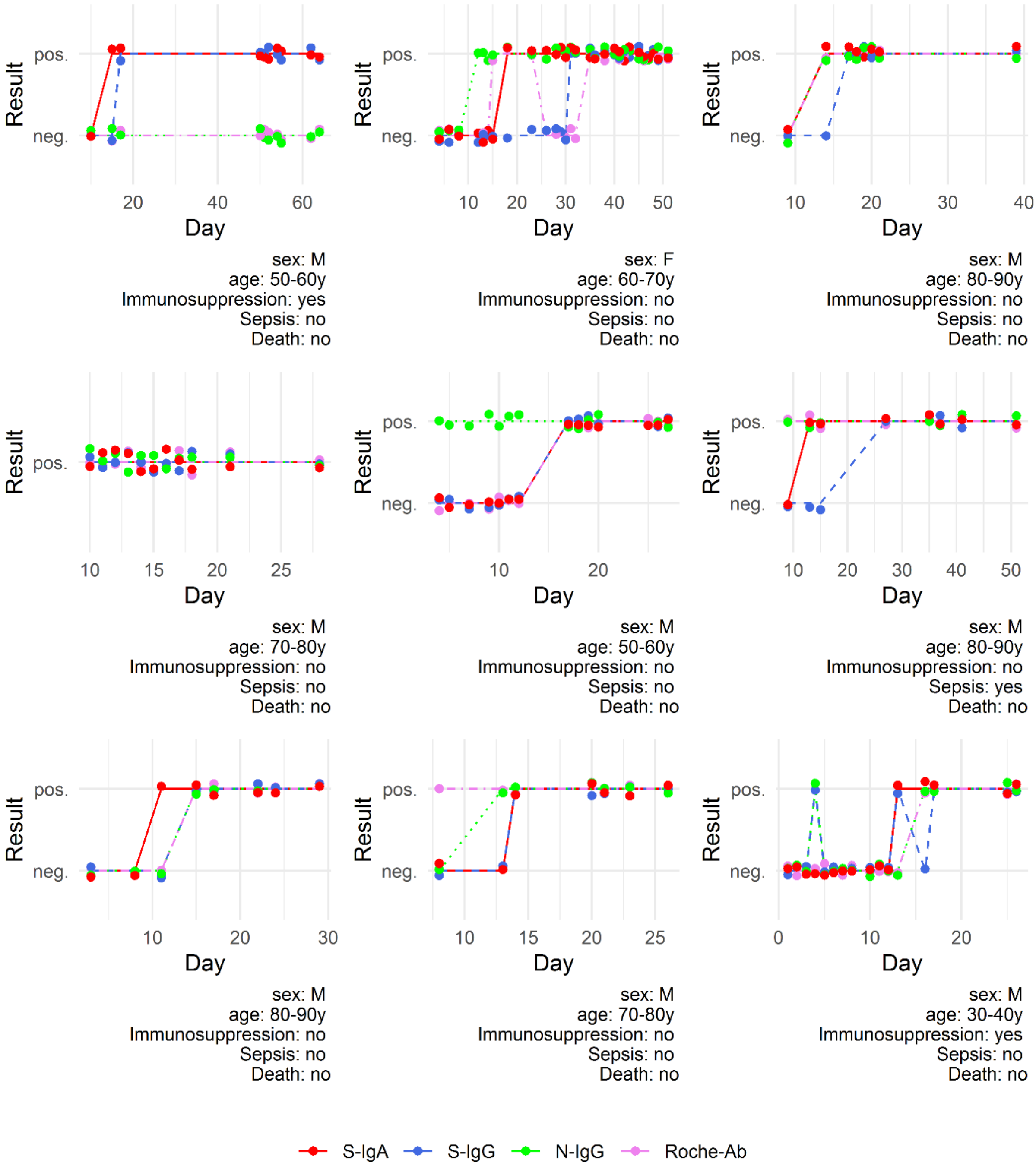


**S8 Fig. Individual qualitative results in the PCR-positive clinical cohort for the four different immunoassays in the non-ARDS group.**
