## Supplementary material for "SARS-CoV-2 antibody immunoassays in serial samples reveal earlier seroconversion in acutely ill COVID-19 patients developing ARDS": S11 Fig

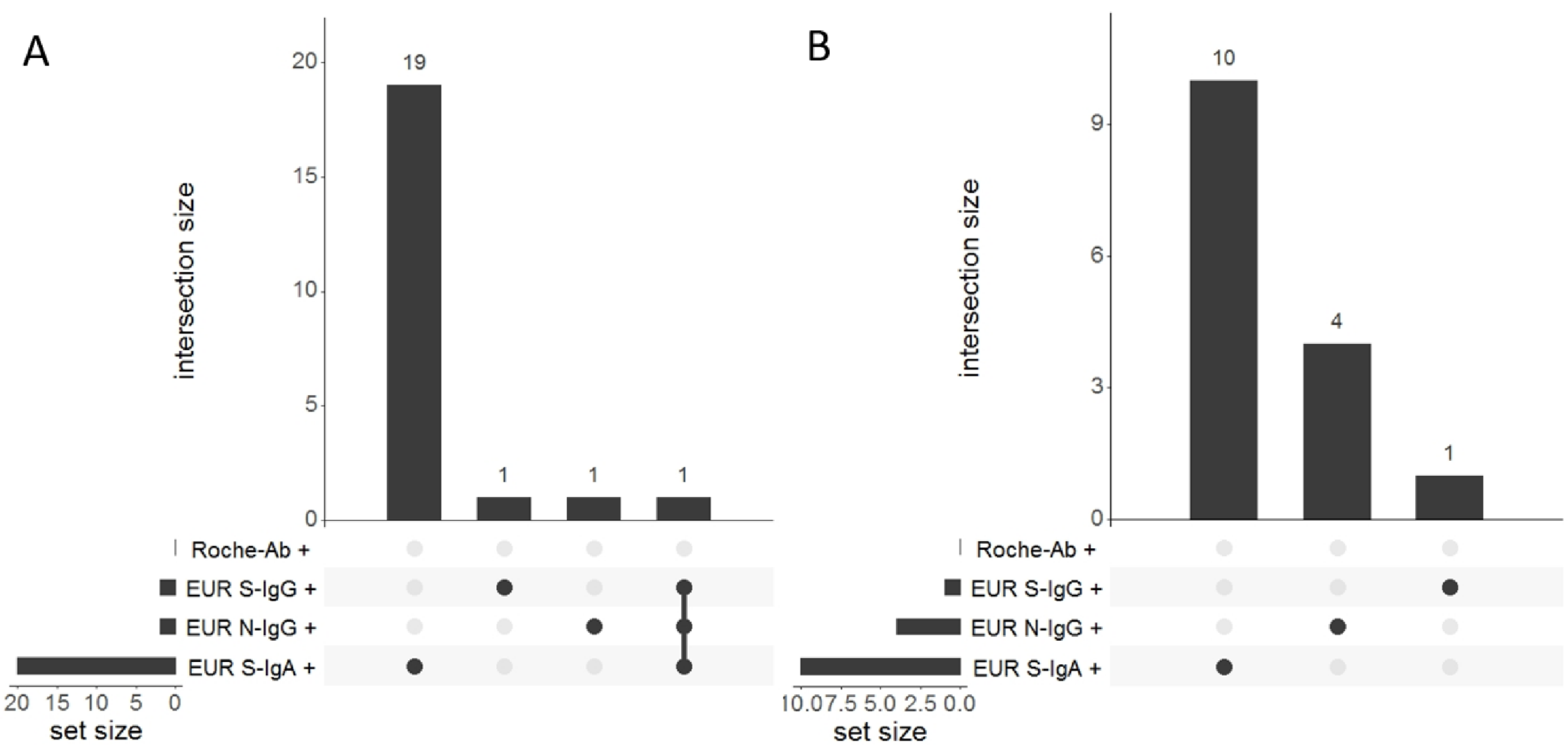


**S11 Fig.** Overlap of positive results between immunoassays in the two negative cohorts. (A) Pre-COVID-19 cohort. (B) PCR-negative clinical cohort.

(This plot was generated using the UpSetR R package) (1)

References

1. Jake R Conway, Alexander Lex, Nils Gehlenborg UpSetR: An R Package for the Visualization of Intersecting Sets and their Properties
doi: https://doi.org/10.1093/bioinformatics/btx364
