## Supplementary material for "SARS-CoV-2 antibody immunoassays in serial samples reveal earlier seroconversion in acutely ill COVID-19 patients developing ARDS": S12 Fig

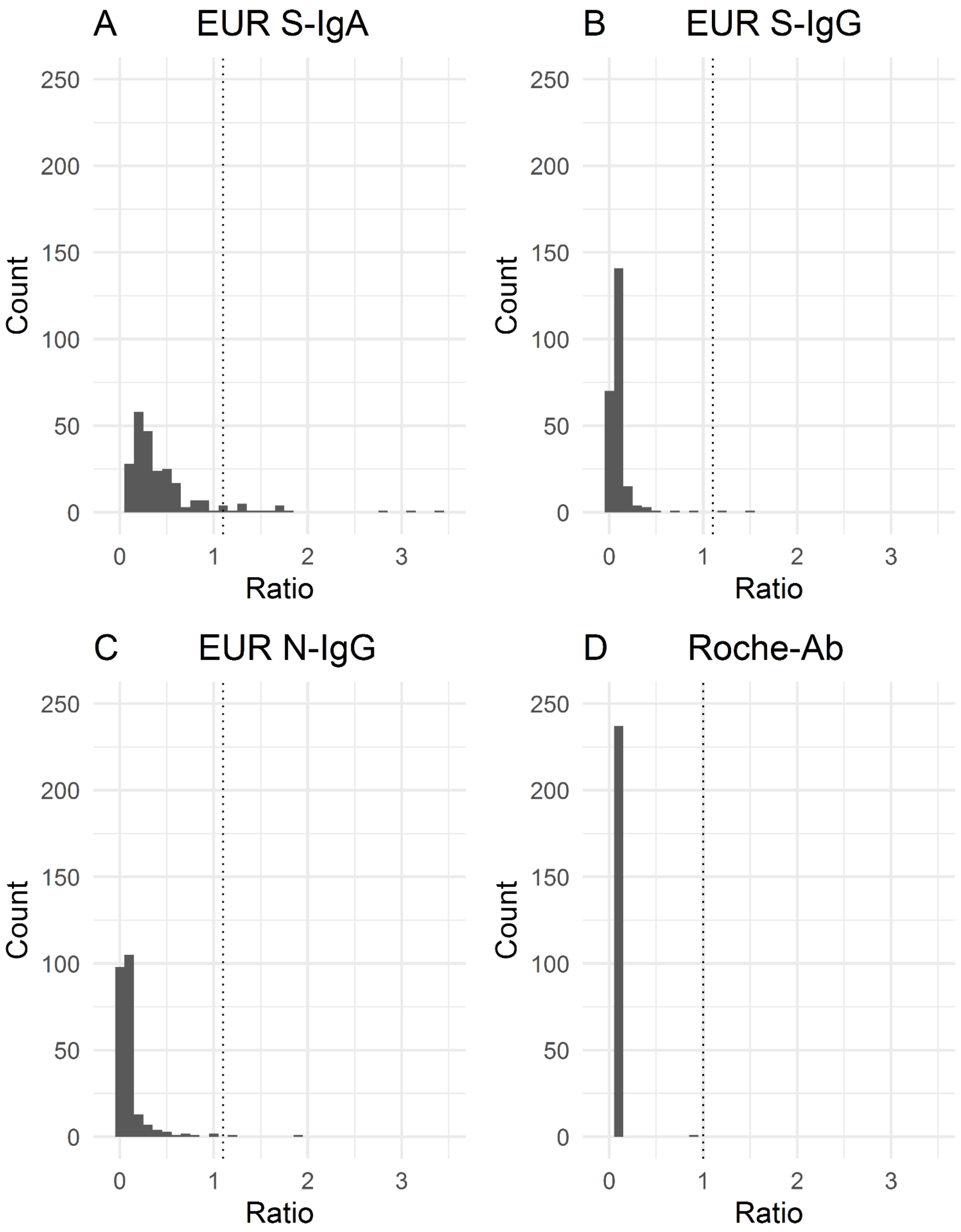


**S12 Fig. Distributions of signal ratios for the four different immunoassays in the pre-COVID-19 cohort.** The dotted lines represent the cutoff values for a positive test result. (A) EUR S-IgA. (B) EUR S-IgG. (C) EUR N-IgG. (D) Roche-Ab.
