## Supplementary material for "SARS-CoV-2 antibody immunoassays in serial samples reveal earlier seroconversion in acutely ill COVID-19 patients developing ARDS": S2 Table

**S2 Table. Median age for true negative and false positive subjects in the negative cohorts.**

| assay | test result | median age pre-COVID19 cohort | median age PCR-negative clinical cohort | median age combined | p-value |
|---|---|---|---|---|---|
| **EUR S-IgA** | neg. | 27 | 70 | 49 | 0.0294 |
| | pos. | 28 | 66 | 30 | |
| **EUR S-IgG** | neg. | 27 | 69 | 48 | 0.7660 |
| | pos. | 41 | 73 | 53 | |
| **EUR N-IgG** | neg. | 27 | 69 | 47 | 0.0253 |
| | pos. | 31 | 93 | 89 | |
| **Roche-Ab** | neg. | 27 | 69 | 48 | / |
| | pos. | / | / | / | |
